# Forecasting COVID-19 Spreading in Canada using Deep Learning

**DOI:** 10.1101/2021.05.01.21256447

**Authors:** Fadoua Khennou, Moulay A. Akhloufi

## Abstract

The novel coronavirus disease 2019 (COVID-19) is disrupting all aspects of our lives as the global spread of the virus continues. In this difficult period, various research projects are taking place to study and analyse the dynamics of the pandemic. In the present work, we firstly present a deep overview of the main forecasting models to predict the new cases of COVID-19. In this context, we focus on univariate time series models in order to analyze the dynamic change of this pandemic through time. We secondly shed light on multivariate time series forecasting models using weather and daily tests data, to study the impact of exogenous features on the progression of COVID-19. In the final stage of this paper, we present our proposed approach based on LSTM and GRU ensemble learning model and evaluate the results using the MAE, RMSE and MAPE for the prediction of new cases. The results of our experiments using the Canadian dataset show that the ensemble model performs well in comparison to other models. In addition, this research provides us with a new outcome regarding the dynamic correlation between temperature, humidity and daily test data and its impact on the new contaminated cases.

## 1 Introduction

The COVID-19 pandemic has spread at a phenomenal rate. From December 2019 to December 2020, 82,872,473 cases have been confirmed, 1,826,809 are reported dead and 60,282,421 recovered worldwide.

Research models that have been proposed since the announcement of the first cases of COVID-19 have made it possible to estimate the size of the epidemic and the severity of the disease. Yet, several questions still remain unanswered due to the new and different characteristics of this virus. While these research studies focus mainly on analyzing the virus behavior, obtaining its related data requires more efforts from multiple stakeholders around the world, which remain difficult to conduct during a large, emerging epidemic.

Therefore, to understand the dynamics of the COVID-19 pandemic, it is necessary to look at the time scale of action of the virus. In this serious situation, many countries report the details of daily infection, hospitalization and mortality indicators.

There are many research projects that are actively working on the forecast of the COVID-19 spread, but only few focused on considering non-patient related risk factors, which can play a direct or indirect role in this spread. The majority of the proposed studies shed light on univariate time series modeling [1, 2, 3]. Furthermore, after an in-depth analysis of recent research predictions, it is noted that only few of them presented merely accurate estimations, this can be explained by the lack of data at the beginning of the outbreak.

The main goal here is to demonstrate the possibility of using myriad modeling strategies to analyze and forecast the future of the COVID-19. The main contributions of this work are as follows:

- Present a broad overview of recent research on COVID-19 forecasting using both statistical and deep learning models.
- Implement univariate and multivariate time series using both statistical and deep learning models.
- Implement an ensemble learning multivariate model based GRU and LSTM for the COVID-19 forecast.

## 2 Related work

Different research projects are actually taking place to forecast the spread of the COVID-19 [4]. While we are in the beginning of the vaccination stage, it is still important to deeply analyse the virus data in order to overcome future viruses and global pandemics. In fact, this presents a significant challenge for researchers specifically in data science, because of the limited data and the unknown characteristics.

Simpler models provide less optimal forecasts because they cannot capture complex patterns and other time-scale changing parameters of infectious disease spread. In addition to that, we notice that this virus has different kind of impact on distinct countries.

The researchers in [5] presented a SEIR based model. Their prediction was effective for China use case in late February, yet they based their study on the 2003 SARS data, to predict the epidemic. And as till now we do not have much current information on the behavior of COVID-19, it remains difficult to validate the used approach. Other researchers [6] proposed a hybrid model for the forecast of new COVID-19 cases, combining ARIMA and wavelet-based forecasting techniques. Their model results emphasized on the deficiencies of the classical ARIMA time series model.

Another research was proposed as an automated real-time forecasting model for COVID-19 transmission using LSTM networks [3]. They used collected data from Johns Hopkins University and Canadian Health authority, provided with the number of confirmed cases until March 31, 2020. In their LSTM model, they trained and tested the network on Canadian dataset; the RMSE error was 34.83 with an accuracy of 93.4% for short term predictions in Canada.

In Brazil, the researchers [7] focused on presenting a forecast model based on CUBIST, RF, RIDGE, SVR, and stacking-ensemble learning models. In their experiment, 180 scenarios (10 datasets, 3 forecasting horizons, and 6 models) were evaluated for the task of forecasting cumulative COVID-19 cases. As a results, SVR and stacking-ensemble learning model were the most suitable tools to forecast COVID-19 cases.

In India, the researchers [8] proposed a data driven model based on LSTM and curve fitting up to 4th April 2020. They compared the results based on the values of the transmission rate r=2.3 before lockdown and r=0.15 after lockdown. They observed that preventive measures have worked well in containing this virus contagion.

Some research studies were conducted in the early stages of the virus spread [9]. At that time, very few data were available, mainly related to China confirmed cases. In this context, the researchers used an Adaptive Neuro-Fuzzy Inference System (ANFIS), which is widely applied in time series prediction and forecasting problems. They used an improved ANFIS model based on a modified flower pollination algorithm (FPA) using the salp swarm algorithm (SSA).

A recent comparative study was conducted by [10]. They implemented ARIMA, LSTM and NARNN on data from Denmark, Belgium, Germany, France, United Kingdom, Finland, Switzerland and Turkey. Their LSTM model showed high performances compared to other models. Yet, they base their study on univariate time series data without considering other external features. Through the analysis of the aforementioned studies related to COVID-19 forecast, we sum up that they focused mainly on using univariate time series modeling or classical statistical models. Our goal is to overcome this shortcoming by developing an ensemble model of GRU and LSTM using external features such as weather data and the number of daily tests for the forecast of new cases in Canada.

## 3 Materials and methods

### 3.1 Dataset

In this study, we used three datasets. We collected weather data from the climate data website, it is an information portal that enables Canadians to access and analyze climate data freely, and provides related information and tools to support adaptation planning and decision-making [11].

We also used the international historical data from the Johns Hopkins University [12], which includes daily coronavirus confirmed, recovered and death cases. We scrapped and processed the data for the Canadian case study.

We finally collected the daily COVID-19 tests data from Our World in Data team repository [13].

### 3.2 Statistical models

As previously noted, different statistical models were implemented for the forecast of COVID-19 [14, 5]. In fact, it has been proven through many studies that classical models achieved better results than sophisticated deep learning methods [15], yet this assumption cannot be generalized for every time series problem. Indeed, the size of data, the seasonality and type of the time series have a tremendous impact on the forecast of future data.

In this study, we focus on implementing both deep learning and statistical models, then evaluating the results depending on varied criteria.

#### 3.2.1 Autoregressive integrated moving average (ARIMA)

There are two categories of models to account for a time series. The first considers that data is a function of time *y* = *f* (*t*).

This category of model can be fitted by the least squares method, or other iterative methods. Fourier transform model analysis is a sophisticated version of this type of model. A second category of models seeks to determine each value of the series as a function of the values which precede it as in eq.(1) :

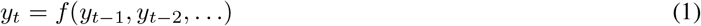

The estimation of ARIMA models assumes that we are working on a stationary series [16]. This means that the series mean is constant over time, as is the variance. The best method to eliminate any tendency is to differentiate, that is, to replace the original series with the series of adjacent differences. A time series that needs to be differentiated to achieve stationarity is considered an integrated version of a stationary series.

Autoregressive processes mean that each point can be predicted by the weighted of a set of previous points, plus a random error term. The model estimates a dependent variable *y*_*t*_ at any timing based on some number of lagged observations.

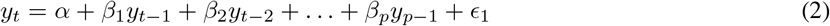

Where the *y*_*t-*1_ is the lag of the time series, *β* is the coefficient of the lag and *α* is the intercept term.

The integration process assumes that each point has a constant difference from the previous point. Moving average processes assume that each point is a function of the errors in the preceding points, plus its own error. An ARIMA model is labeled as ARIMA model (p, d, q), in which:

- p is the number of auto-regressive terms
- d is the number of differences
- q is the number of moving averages

We declare a process (*x*_*t*_) where t ∈ ℕ is an ARIMA process with order (p,d,q), as ARIMA (p,d,q) if:

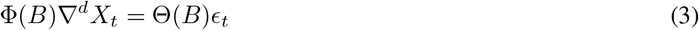

Where:

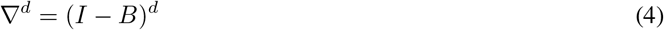

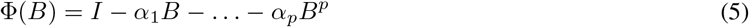

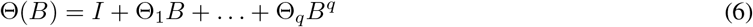

#### 3.2.2 Seasonal Autoregressive Integrated Moving Average (SARIMA)

One of the major problems with an ARIMA is that it does not support seasonality. The autoregressive seasonal integrated moving average (SARIMA), is an extension of ARIMA that explicitly supports the seasonal component [17]. Seasonality itself is treated as an ARIMA, with terms similar to the non-seasonal components of the model. However, it implies to be based on lags equivalent to the periodicity of the season.

There are four seasonal parameters that are not part of ARIMA, which must be configured. The SARIMA process is presents as: SARIMA(p,d,q)(P,D,Q)m where:

- P: Seasonal autoregressive order
- D: Seasonal difference order
- Q: Seasonal moving average order
- m: The number of time steps for a single seasonal period

While SARIMA and ARIMA models are only taking into account one parameter for the time series forecast, it is important to note that sometimes forecasting the future based solely on time cannot be very efficient. Thus, we are going to need a lot more outside data features and factors in order to start predicting a variable. In fact, ARIMA performs well when working with a time series where the data is directly related to the time stamp. In this context, SARIMA provide a new model called SARIMAX with exogenous variable.

### 3.3 Deep learning models

#### 3.3.1 Recurrent neural network (RNN)

A recurrent neural network (RNN) is a network of artificial neurons with recurrent connections, that is, interconnected neurons that can cycle. These networks have proven their effectiveness in modeling problems of automatic recognition of speech, handwriting, computer vision and in time series forecasting.

In RNN, the output of any layer depends not only on the current input but also on the set of previously reported inputs.

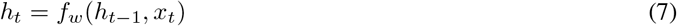

As presented in eq.(7), this unique function gives it a significant benefit over other neural networks by taking advantage of previously collected inputs estimating outputs at the later level.

#### 3.3.2 Long Short Term Memory (LSTM)

Recurrent neural networks are widely used in applications such as automated language processing. However, they have learning problems when the processed sequences become too long. The gradient may become too weak at the end of the chain. In this vein, LSTM networks help solve this problem.

Both LSTM and GRU were created as a method to efficiently manage short and long term memory through their gate systems. If there are many variations, the original versions are still very widely used in the best deep learning models for automatic natural language processing.

LSTM, which stands for Long Short-Term Memory [18], is a cell made up of three gates: these are calculation areas that regulate the flow of information (by performing specific actions). There are also two types of outputs (called states), the LSTM architecture is schematized in figure 1.

**Figure 1:**
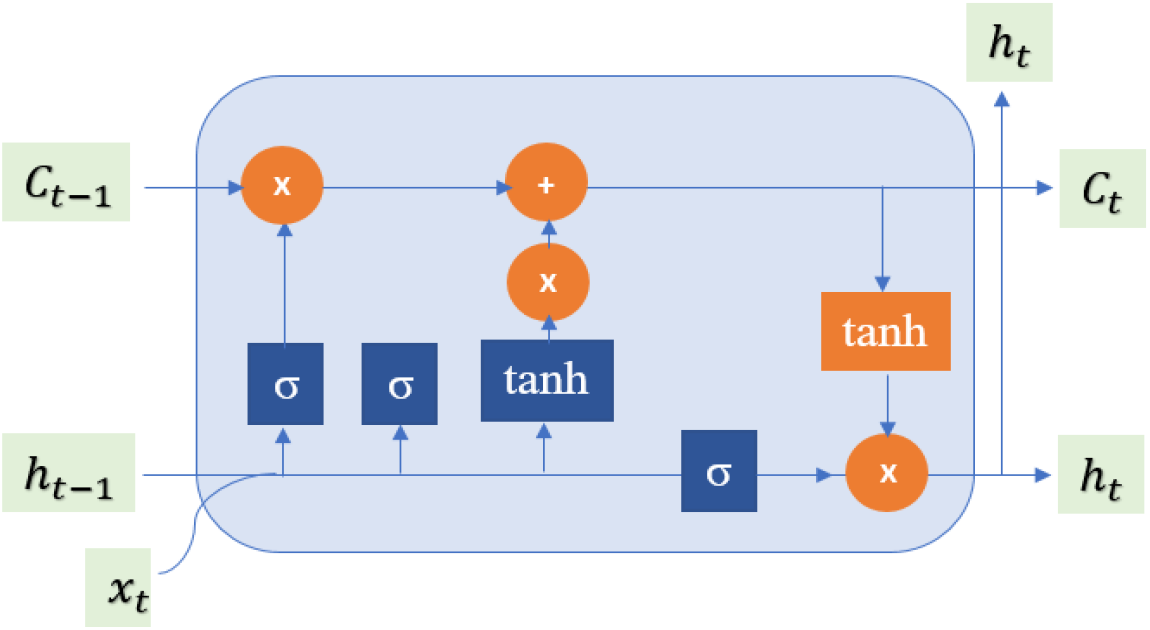
LSTM architecture

All recurrent neural networks have the form of a chain of recurrent modules of neural network. The gates contain sigmoid activations, which compresses values between 0 and 1. That is helpful to alter or forget data because any number getting multiplied by 0 is 0, will cause values to disappears or be “forgotten.”

#### 3.3.3 Gated recurrent unit (GRU)

The Gated Recurrent Units were initially proposed by [19]. Using the update and reset gates, GRU solves the gradient vanishing problem faced in RNNs. As described in figure 2, at each time t, we have the state h and the current time input x. The reset gate discovers which of the input data will be overlooked. For example, in this paper, we use weather data to forecast the number of confirmed cases, in this case, we may have some element in the input like humidity, which the network would know to be unrelated to the COVID-19 forecast and “reset it.”

**Figure 2:**
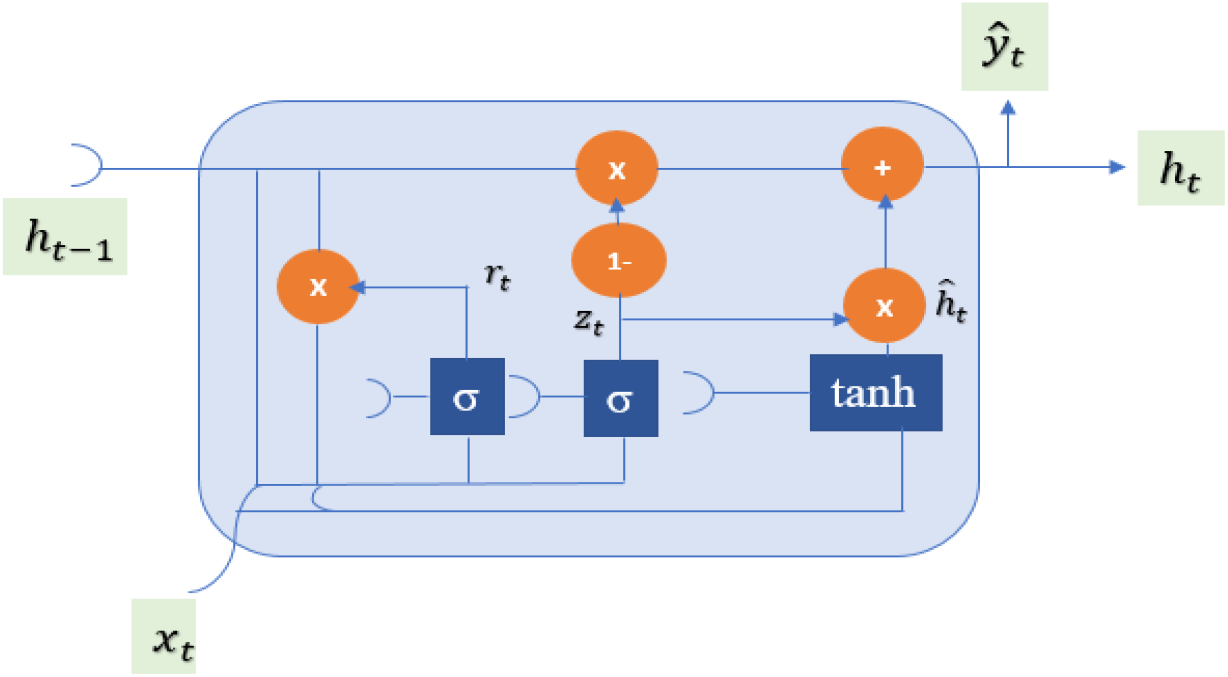
GRU architecture

The upgrade gate knows what data to upgrade from the input with newer data in the environment. In our case study, there might be some data such as humidity that the network may learn to update or change the previous value with the newer one in the input.

### 3.4 Forecasting COVID-19 cases

In this paper, we focus on two main steps to train the models. The first concerns univariate time series modeling to estimate the forecast for future cases of COVID-19 based only on time stamps data, while the second concerns multivariate modeling to estimate the forecast based on different features as temperature, precipitations, humidity and the number of daily tests.

#### 3.4.1 Data preprocessing

The collected weather data (mean temperature, humidity and precipitations) was merged with the database containing the number of confirmed cases, the number of deaths, the number of daily tests as well as the total number of tests for each province in Canada. We did not process recovered cases data since it was missing for most provinces.

#### 3.4.2 Proposed approach

This study introduces deep learning and statistics modelling for the forecast of COVID-19 in Canada. The proposed methodology consists of the main following steps described in figure 3.

**Figure 3:**
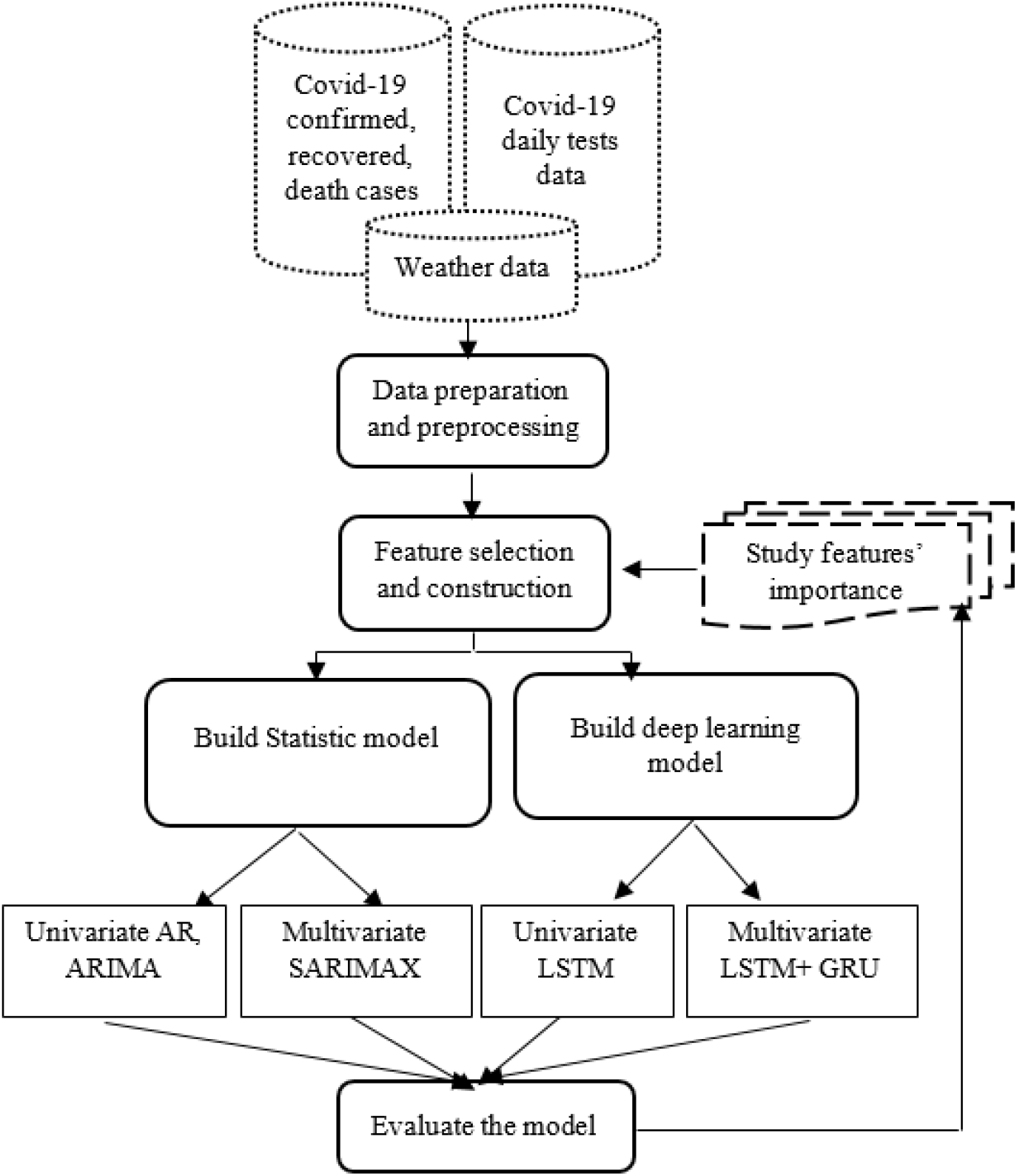
Proposed approach

1. Collecting and gathering data.
2. Preprocessing and preparing data.
3. Conducting feature selection and extraction
4. Implementing univariate and multivariate time series models, using both statistical and deep learning models.

In order to get better results in forecasting COVID-19 confirmed cases, we used model averaging. For both LSTM and GRU models, we created one single layer with 100 neurons and a dense layer to consolidate the output. We then used Adam optimizer for both models. Taking into account that ensemble learning is considered the best way to get the most accurate results without overfitting, while having minimum loss, we enhanced the previous methods using an ensemble of both algorithms using the average ensembling technique by calculating the average of the models’ predictions.

#### 3.4.3 Evaluation metrics

The performance of the proposed approach is evaluated based on three performance measures : MAE, RMSE and MAPE.

The mean absolute error (MAE) represents the difference between the actual and the measured value. By evaluating this metric, we can get an idea of the precision of a value. Indeed, if we know the measured value and the real value, it is then possible to perform a simple subtraction to find the absolute error. This is obtained using eq.(8).

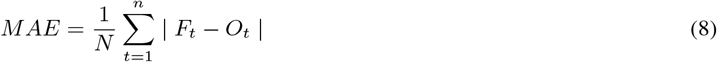

This root mean squared error (RMSE) is a statistical measure, which calculates the average magnitude of errors. It does not indicate their direction. In fact, the mean square error gives more weight to large errors than to others when calculating the mean, compared to the mean absolute error. So, when the root mean square error is much greater than the mean absolute error, it means an increased frequency of large errors. It is therefore an appropriate statistical when large errors are very undesirable. The RMSE is calculated using eq.(9).

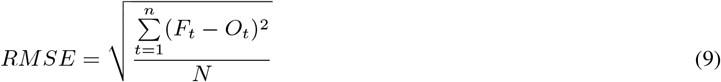

The mean absolute percentage error (MAPE) helps to depict the difference between an exact value and an estimated value. It is used generally to compare the fitted values of different time series models. The MAPE is calculated using eq.(10).

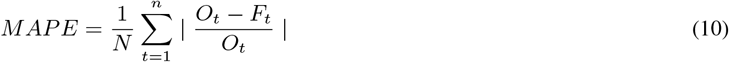

## 4 Results

In this section we analyze the obtained results for the COVID-19 forecast applied to the Canadian case study, using different methods.

### 4.1 Forecasting using univariate time series

In the first phase, we used univariate time series modeling in order to forecast the new cases in Canada without considering any exogenous variable.

For that, we developed three models, the first is a statistical auto regressive model, we forecast using a linear combination of past values of the variable. We split the data into 80% of train data and 20% of test sets, then we trained the model using the TSA python library for statistical modeling [20]. In order to fit our model, we tested over 7 and 8 lags representing the p parameter of the auto regressive model. The forecast results are presented in table 1.

**Table 1:**
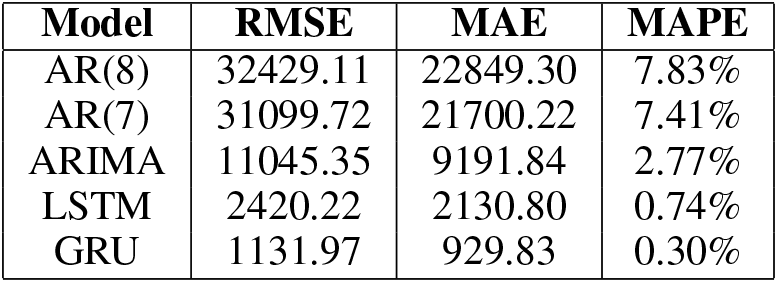
Evaluation metrics results

| Model | RMSE | MAE | MAPE |
| --- | --- | --- | --- |
| AR(8) | 32429.11 | 22849.30 | 7.83% |
| AR(7) | 31099.72 | 21700.22 | 7.41% |
| ARIMA | 11045.35 | 9191.84 | 2.77% |
| LSTM | 2420.22 | 2130.80 | 0.74% |
| GRU | 1131.97 | 929.83 | 0.30% |

The second is the ARIMA model, we used the same number of training and testing sets for each of the three models. Here we used the auto ARIMA function to obtain the best combination of the (p, d, q) orders. This is schematized in figure 4.

**Figure 4:**
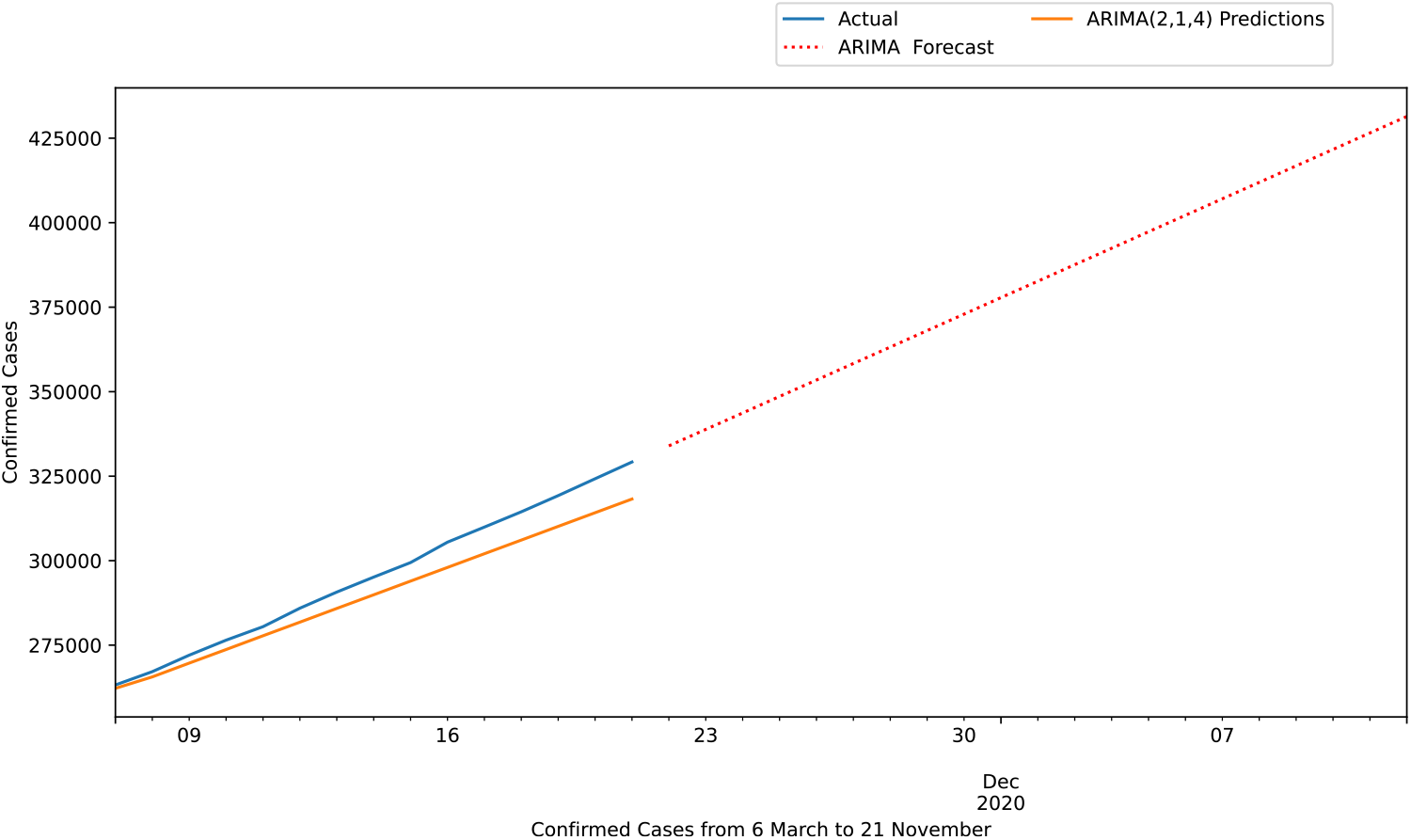
Forecasting using ARIMA

The third is a deep learning model based on LSTM and GRU. Our network is composed of a first layer with 100 neurons and a dense node to consolidate the output. Adam was used as optimizer. We used the time series generator class to create batches of temporal data. We selected a validation set to avoid over-fitting and tested over 20 time steps to predict the next data point. We also added the “restore best weights” callback, which saves the best model observed during training after 20 iterations. Table 1 illustrates the results of the forecast.

Based on data from all provinces, LSTM and GRU achieved better results for the forecast of new cases. With a RMSE of 2420.22 and 1131.97 respectively. As for testing over different AR lags, the AR(7) provided the best results in the autoregressive modeling category. While we notice that statistical models provide promising results for COVID-19 forecast, it is crucial to note that they have some limitation in regards to model tuning and optimization. In the LSTM model we used the early stopping function to avoid overfitting due to too many iterations and consolidated a dropout function for a better generalization.

For Stacked, Bi-directional and convolutional LSTM the researchers [21] obtained a MAPE of 4%, 3.33% and 2.17% for INDIA. As for USA, they obtained 10%, 6.66% and 2% respectively. Other researchers [22] compared the results of (ITALY, JAPAN, SPAIN, UK and US) using both LSTM and ARIMA. For ARIMA their MAPE ranges between 1.36% and 21.75% and for LSTM it ranges between 4.39% and 16.67%. It is important to note that, since each country has a glaring difference in the number of cases, it is difficult to compare the results, as they depend mainly on the used data. Yet, with a very high error values, it clearly impacts heavily the prediction of the models. That is to say, it is crucial to focus deeply on each country, at first, so as to understand the impacts of different variables on the progression of the virus. In the next section, we will focus on modeling using multivariate time series for Canadian provinces, where the disease progresses differently.

### 4.2 Forecasting using multivariate time series

The difference between a univariate and a multivariate time series is the number of values at each instant of time. These multiple values interact with each other and give us a clear idea of how they impact the predicted value in a specific time.

In order to predict COVID-19 cases using multivariate time series, we firstly assessed parametric and non parametric associations and correlations between all variables, identified outliers and imputed missing values. We present in figure 5 the forecasting models for four Canadian provinces with high number of confirmed cases. In this vein, the model performs well as training and validation loss are projecting at a minimal value. The curves are getting flattened as the number of epochs increases. After different iterations, we fine tuned the model using a batch size of 4, 20 lag timestamps and final features of (humidity, mean temperature and number of daily tests).

**Figure 5:**
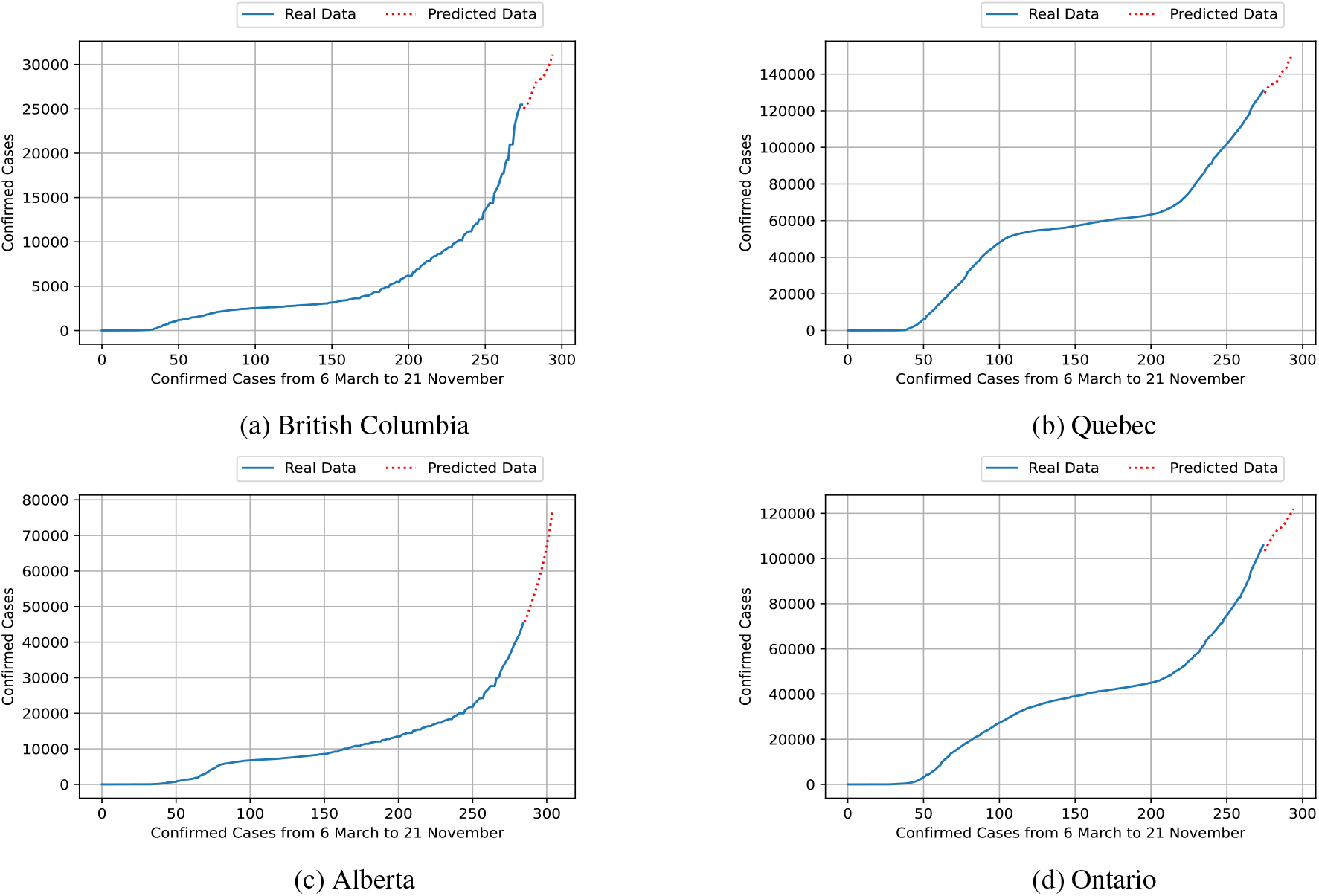
Forecasting new cases for the next 20 days

For Quebec and Ontario as presented in figure 5b and figure 5d, we can see that the forecast for the next 20 days (starting from November,21 2020) is reaching more than 140000 and 120000 cases respectively. As for Alberta, figure 5c shows an highly exponential increase reaching 70000, while for British Columbia it reaches more than 30000. Table 2 presents a summary of the forecasting errors for the four models tested on all the Canadian provinces.

**Table 2:**
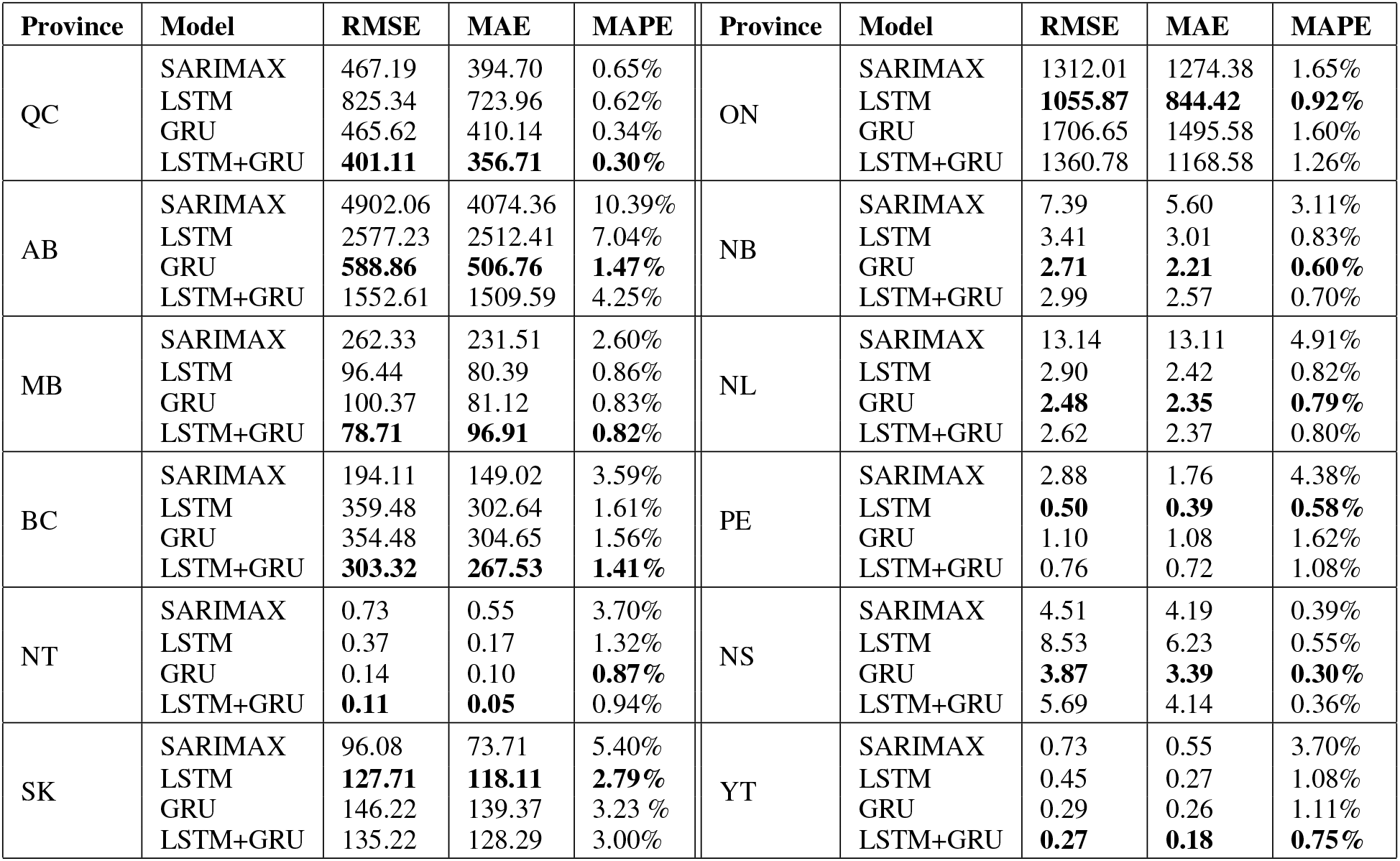
Evaluation metrics of multivariate time series models

| Province | Model | RMSE | MAE | MAPE | Province | Model | RMSE | MAE | MAPE |
| --- | --- | --- | --- | --- | --- | --- | --- | --- | --- |
| QC | SARIMAX | 467.19 | 394.70 | 0.65% | ON | SARIMAX | 1312.01 | 1274.38 | 1.65% |
|  | LSTM | 825.34 | 723.96 | 0.62% |  | LSTM | **1055.87** | **844.42** | **0.92%** |
|  | GRU | 465.62 | 410.14 | 0.34% |  | GRU | 1706.65 | 1495.58 | 1.60% |
|  | LSTM+GRU | **401.11** | **356.71** | **0.30%** |  | LSTM+GRU | 1360.78 | 1168.58 | 1.26% |
| AB | SARIMAX | 4902.06 | 4074.36 | 10.39% | NB | SARIMAX | 7.39 | 5.60 | 3.11% |
|  | LSTM | 2577.23 | 2512.41 | 7.04% |  | LSTM | 3.41 | 3.01 | 0.83% |
|  | GRU | **588.86** | **506.76** | **1.47%** |  | GRU | **2.71** | **2.21** | **0.60%** |
|  | LSTM+GRU | 1552.61 | 1509.59 | 4.25% |  | LSTM+GRU | 2.99 | 2.57 | 0.70% |
| MB | SARIMAX | 262.33 | 231.51 | 2.60% | NL | SARIMAX | 13.14 | 13.11 | 4.91% |
|  | LSTM | 96.44 | 80.39 | 0.86% |  | LSTM | 2.90 | 2.42 | 0.82% |
|  | GRU | 100.37 | 81.12 | 0.83% |  | GRU | **2.48** | **2.35** | **0.79%** |
|  | LSTM+GRU | **78.71** | **96.91** | **0.82%** |  | LSTM+GRU | 2.62 | 2.37 | 0.80% |
| BC | SARIMAX | 194.11 | 149.02 | 3.59% | PE | SARIMAX | 2.88 | 1.76 | 4.38% |
|  | LSTM | 359.48 | 302.64 | 1.61% |  | LSTM | **0.50** | **0.39** | **0.58%** |
|  | GRU | 354.48 | 304.65 | 1.56% |  | GRU | 1.10 | 1.08 | 1.62% |
|  | LSTM+GRU | **303.32** | **267.53** | **1.41%** |  | LSTM+GRU | 0.76 | 0.72 | 1.08% |
| NT | SARIMAX | 0.73 | 0.55 | 3.70% | NS | SARIMAX | 4.51 | 4.19 | 0.39% |
|  | LSTM | 0.37 | 0.17 | 1.32% |  | LSTM | 8.53 | 6.23 | 0.55% |
|  | GRU | 0.14 | 0.10 | **0.87%** |  | GRU | **3.87** | **3.39** | **0.30%** |
|  | LSTM+GRU | **0.11** | **0.05** | 0.94% |  | LSTM+GRU | 5.69 | 4.14 | 0.36% |
| SK | SARIMAX | 96.08 | 73.71 | 5.40% | YT | SARIMAX | 0.73 | 0.55 | 3.70% |
|  | LSTM | **127.71** | **118.11** | **2.79%** |  | LSTM | 0.45 | 0.27 | 1.08% |
|  | GRU | 146.22 | 139.37 | 3.23 % |  | GRU | 0.29 | 0.26 | 1.11% |
|  | LSTM+GRU | 135.22 | 128.29 | 3.00% |  | LSTM+GRU | **0.27** | **0.18** | **0.75%** |

We note that LSTM did not provide good results in comparison to the GRU model. In fact, GRU presented the best MAPE, RMSE and MAE for Alberta, Nova Scotia, Newfoundland and Labrador and New Brunswick, while LSTM delivered good performance for Saskatchewan, Ontario and Prince Edward Island. The ensemble of GRU and LSTM provided the best results for the rest of the provinces; Yukon, Quebec, Manitoba, British Columbia, Ontario and Northwest Territories. In fact, it is proven that LSTM works best when having more data, while GRU is less parametric and boosted the results for the ensemble model.

The SARIMAX model achieved very low results compared to other models. This is mainly affected by the absence of clear seasonality in the majority of the provinces. In addition, after altering between different exogenous features, the number of daily tests proved to be the only variable that has high correlation with the number of confirmed cases for this statistical model.

### 4.3 Discussions and analysis

The forecast for Quebec, which has the highest number of cases, demonstrates the correlation between temperature, humidity and the actual number of cases, with a RMSE of 401.11 and a MAPE of 0.30%.

In addition to Quebec, Alberta and Ontario also captured the association between the above mentioned features with a RMSE of 588.86, 1055.87 and a MAPE of 1.47%, 0.92% respectively.

Overall, this study presented a contribution in the COVID-19 research by providing a comparison between univariate and multivariate time series forecast. This research highlights the potential of ensemble modelling and GRU in forecasting new cases. On the one hand, implementing the models on data for all Canadian provinces gave us the ability to understand how the models perform on varied portion of data.

On the other hand, the use of deep learning models add relevant value in optimizing the performance of the models compared to statistical models. As such an approach based upon ensembling LSTM and GRU, the model is well suited to extension via the inclusion of additional new data for the COVID-19 or for other countries. One major limitation of this research is related to the data availability. Once we get the opportunity to capture more data we can add more features and enhance its performance as a generalisation tool for the SARS-CoV-2.

## 5 Conclusion

In this work, we proposed an ensemble learning model based on LSTM and GRU to predict new cases using Canadian datasets.

The results depicted apparent correlation between humidity, temperature and the number of confirmed cases in provinces with high cases such as Quebec, Alberta and Ontario. The performance for each model has been verified using RMSE, MAE and MAPE.

Further studies need to be conducted in our future work to study new features that may have a direct impact on the forecast. An example of socioeconomic and demographic data can further be considered.

## Data Availability

The data used are available publicly as described in the dataset section.

## Ethical conduct of research

No IRB/oversight body approval or exemption was necessary as the data is publicly available.

## Availability of Data

The data used are available publicly as described in the dataset section.

